# Alu overexpression leads to increased double-stranded RNA in dermatomyositis

**DOI:** 10.1101/2024.11.02.24316621

**Authors:** Rayan Najjar, Andrew Mammen, Tomas Mustelin

## Abstract

Dermatomyositis is an autoimmune condition characterized by a high interferon signature of unknown etiology. Because genes constitute only <2% of our genomes, there is a need to explore the role of the non-coding genome in disease pathogenesis. Our genomes include roughly 1.2 million Alu elements occupying about 10% of the genome and can form double-stranded (ds)RNA capable of triggering MDA5 leading to interferon production. We aligned muscle biopsy RNA sequencing data to the Telomere-to-Telomere reference genome and quantified short interspersed elements including Alus. Dermatomyositis muscle (n=39) showed a global elevation in Alu expression as well as an increased expression of unique Alu elements (n=557, p<0.05) compared to healthy controls (n=34), in a pattern not seen in other myositis types (n=81). The majority (75.3%) of these Alus originated from genomic regions outside genes, with a hot spot of expression on chromosome 19. A subset of the uniquely overexpressed Alus (n=167) correlated strongly with interferon stimulated genes and markers of myositis activity. Since Alu transcripts have a propensity to form dsRNA and are the major targets of both ADAR and MDA5, we quantified the A-to-I RNA editomes inside Alus and found a uniquely expanded Alu editome in dermatomyositis compared to other myositis types, reflecting an increase in dsRNA. Edited Alus clustered on chromosome 19, which is known to have the highest concentration of dsRNA. We hypothesize that overexpressed Alus in dermatomyositis form endogenous dsRNA that exceed the capacity of RNA editing enzymes and trigger dsRNA sensors leading to interferon production.

## INTRODUCTION

Idiopathic inflammatory myopathies (here referred to as “myositis”) include multiple clinical entities: dermatomyositis (DM), antisynthetase syndrome (AS), immune-mediated necrotizing myopathy (IMNM), inclusion body myositis (IBM), and others. They are characterized by upregulation of interferon stimulated genes (ISGs)[1] and the presence of specific autoantibodies,[2] but their etiologies remain unknown.

Short interspersed elements (SINEs) are a subclass of genomic retroelements including two main types: Alu elements and mammalian-wide interspersed repeats (MIRs). Alu elements are prevalent in the human genome with over 1.2 million copies occupying approximately 10% of the human genome, while MIRs constitute about 2.6% of the human genome.[3, 4] Alu elements are approximately 300 bp in length and are composed of similar left and right arms, with an internal polyA tail (**Fig. 1a**). The two arms are derived from 7SL, the RNA component of the signal recognition particle (which, in turn, is a target of autoantibodies in one myositis subtype). MIRs are about 260 bp in length and include 3 parts: a tRNA-derived segment, a unique conserved core, and a segment from a long interspersed nuclear element (**Fig. 1b**). Many genes include embedded SINEs inside introns and UTRs.[5] SINEs have two internal promotor sites for RNA polymerase (Pol) III[5] and are transcribed in two ways: through Pol III directed transcription[6, 7] as well as through passive expression during gene transcription by Pol II.

**Fig. 1:**
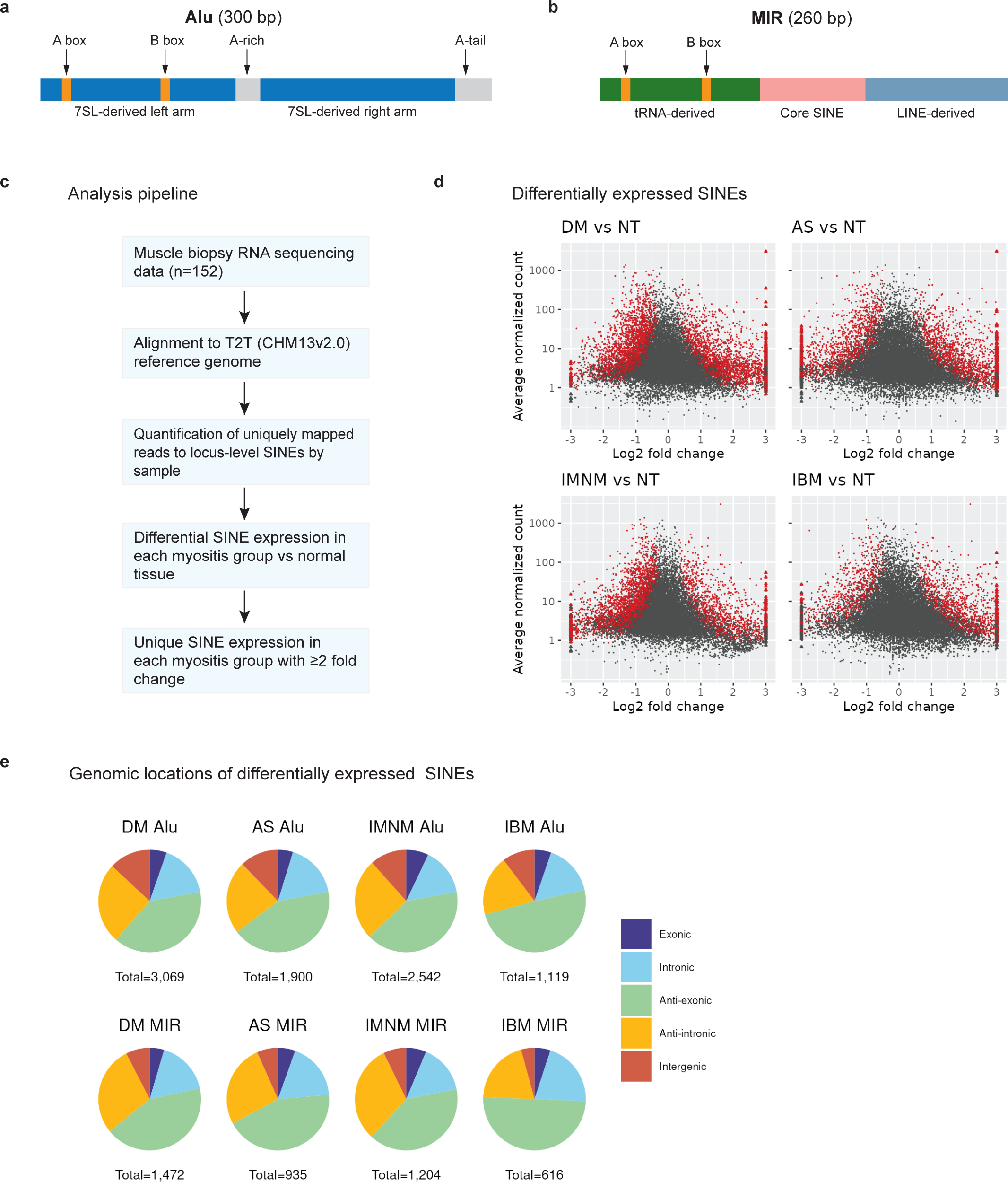
Short interspersed elements (SINEs): structure, study pipeline, and differential expression in muscle tissues from myositis patients. **a**, Schematic illustration of an Alu element. **b**, Schematic illustration of a MIR element. **c**, Our analysis pipeline. **d**, Volcano plots of the differential expression of SINEs in each myositis subtype (DM=dermatomyositis, AS=anti-synthetase syndrome, IMNM=immune-mediated necrotizing myopathy, IBM=inclusion body myositis) compared to normal muscle tissue (NT). **e**, The proportion of SINEs located in exons, introns, the opposite strand of exons or introns, or between genes.

Alus can create double-stranded (ds)RNA leading to interferon production. Due to their sequence similarity and abundance, Alus can form dsRNA when sense and antisense Alu sequences are present in a transcript and anneal to create a dsRNA loop structure.[8] Individual Alu transcripts can also anneal with antisense Alu sequences in other transcripts. A dsRNA structure of sufficient length will activate the dsRNA sensor melanoma differentiation-associated gene 5 (MDA5),[9] which is a target of autoantibodies in one subtype of DM. This leads to downstream signaling to activate interferon production and is prevented under normal conditions by adenosine deaminase RNA specific (ADAR) that catalyzes the post-transcriptional RNA editing of adenosine (A) to inosine (I) in dsRNA, thereby creating breaks disrupting the double-strandedness of RNA and preventing MDA5 triggering. Alus are the major source of dsRNA bound to MDA5[9, 10] including in cancer cells after exposure to demethylating agents where Alus form dsRNA, trigger MDA5, and activate interferon production[11] creating an immunogenic high-interferon viral mimicry state that can be a potent anti-cancer therapeutic.

Through transcriptomic analysis of muscle tissue from myositis patients and healthy controls, we explore the expression of SINEs in myositis types including unique expression in one type not seen in others. Second, we correlate SINEs with ISGs and disease activity markers. And finally, we quantify RNA A-to-I editing to map the editomes in myositis.

## RESULTS

### Expression of SINEs in myositis subtypes

We aligned muscle biopsy RNA-sequencing data (n=152) to the T2T (CHM13v2.0) reference genome,[3] which is the first truly complete human genome allowing more accurate quantification of locus-level expression of SINEs. Analyses were done by myositis subtypes (DM=39, AS=15, IMNM=51, IBM=15) vs normal tissue (n=32). Our analysis pipeline is illustrated in **Fig. 1c**. We found differential expression of thousands of significantly altered SINEs in each myositis subtype vs normal tissue (**Fig. 1d**), highest in DM (n=4,568), and lowest in IBM (n=1,749).

The majority of differentially expressed SINEs were either Alu elements (≈2/3) or MIRs (≈1/3) in each myositis subtype. SINEs are widely distributed in our genome, including in exons and introns of genes. To explore the distribution of differentially expressed SINEs, we mapped their genomic locations, classifying them into the following mutually exclusive groups: exonic, intronic, antisense exonic, antisense intronic, or intergenic. Out of differentially expressed Alus and MIRs by myositis subgroup, 15.1%-20.6% were intronic, 4.6%-7.0% exonic, while the majority (74.4%-78.5%) originated from intergenic or antisense strands to genes (**Fig. 1e**), indicating that these SINEs were not transcribed passively during gene transcription.

### Alus are highly and uniquely expressed in DM

SINEs that are expressed in one myositis subtype and not others are more likely to reflect distinct and more meaningful disease biology than those Alus expressed in all myositis subtypes. Therefore, we identified SINEs with unique expression in each myositis subtype compared to the others, focusing on elements with ≥2-fold change (**Fig. 2a**). DM had a high number of uniquely expressed SINEs (n=740), most of which were overexpressed Alus (577, 78.0%) with a median fold increase of 4.4 (interquartile range [IQR] 2.9-8.8), compared to 53 underexpressed Alus with a median fold decrease of 2.4 (IQR 2.2-2.8) in DM versus controls. This contrasted with the other myositis types with only 39-81 uniquely overexpressed Alus. Therefore, we decided to further characterize and explore the uniquely overexpressed Alus in DM.

**Fig. 2:**
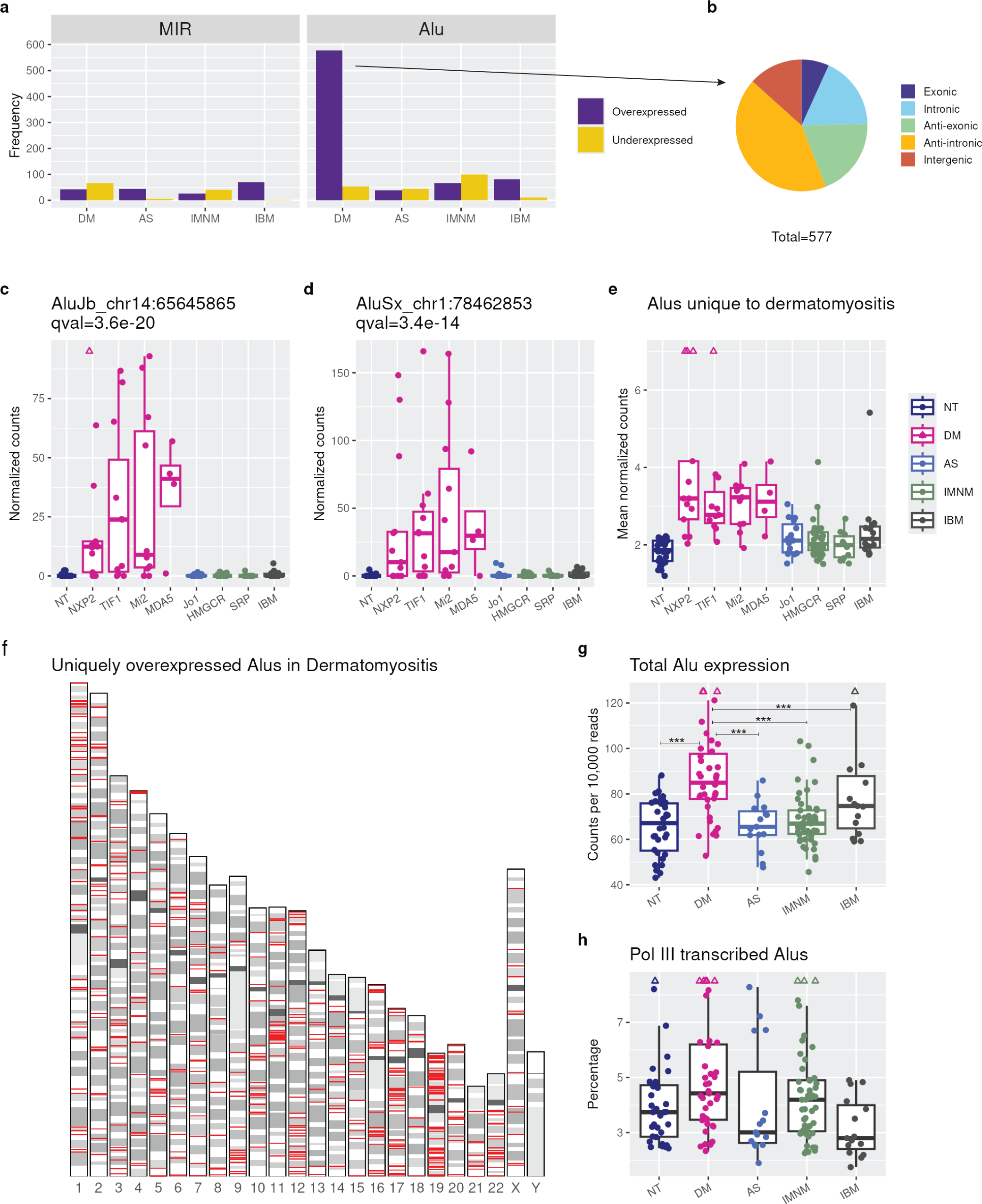
Unique and overall Alu expression in myositis. **a**, The number of MIR and Alu elements expressed ≥2-fold lower or ≥2-fold higher in each myositis subtype compared to healthy muscle. **b**, Genomic location of the 577 uniquely increased Alu elements with respect to genes. **c-e**, The expression levels of the indicated Alu element(s) in myositis subtypes further subdivided by myositis-specific autoantibodies. Adjusted p-values (q values) are shown. **e**, Chromosomal location (red lines, not to scale) of the uniquely overexpressed Alu elements in dermatomyositis. **g**, Total Alu reads in myositis subtypes, Poisson regression, ***= q<0.001. **h**, The percent of Alu transcripts deemed Pol III generated based on low coverage of reads mapping immediately upstream or downstream of the Alu element. For all boxplots, center line, median; box limits, upper and lower quartiles; whiskers, 1.5x interquartile range; points beyond whiskers, outliers (triangles when higher than maximum whisker)

Most of the uniquely overexpressed Alus in DM originated from intergenic/antisense regions (**Fig. 2b**). Still, 39 (6.8%) Alus were in exonic and 103 (17.9%) in intronic regions.

Overexpression of the unique DM Alus was seen similarly in all four subtypes of DM classified by myositis specific autoantibodies (anti-NXP2, anti-TIF1, anti-Mi2, and anti-MDA5). The Alu element with the lowest q-value was an AluJb with the coordinates chr14:65,645,865-65,646,165+ (‘+’ denotes the positive DNA strand), which was uniquely overexpressed in all four subtypes of DM (**Fig. 2c)**. A similar pattern was seen in the second lowest p-value Alu (**Fig. 2d**), and when considering the average expression of all uniquely overexpressed DM Alus (n=577) by sample (**Fig. 2e**).

Unique Alus in DM originated from widespread chromosomal regions, but also included hot spots (**Fig. 2f**). There were 66 (11.4%) unique Alus from chromosome 19, more than double what is expected in this chromosome if the distribution of unique Alus was random among chromosomes (expected 4.8%, p=3.5 x 10^-13^). For comparison, chromosome 1, which is the longest one and harbors 8.6% of all Alus, had 52 (9.0%) of the DM unique Alus. Additionally, chromosome 17 contained 42 (7.3%) of the unique Alus in DM (expected 4.8%, p 0.048). In contrast, chromosome 8 had a lower number (n=11, 1.9%) of unique Alus than expected (4.1%, p 0.010).

To explore if the Alu overexpression in DM is limited to uniquely expressed Alus or a general phenomenon, we quantified the cumulative total RNA-seq coverage of Alus and found an increase in DM (**Fig. 2g**), suggesting a global overexpression.

We next explored transcription of Alus through their own promoters. We considered Alus with low RNA-coverage in the genomic regions upstream and downstream of the Alu body to be RNA polymerase III (Pol III)-transcribed, using established methodology.[6] We applied this to intergenic/antisense Alus, since Alus embedded in genes are highly likely to be transcribed by RNA Polymerase II during gene expression. We discovered 4,347 Pol III-transcribed Alus, which was a low percent of all expressed Alus per sample in myositis groups and healthy controls (median 2.8%-4.4%, **Fig. 2h**). However, the percent of Pol III-transcribed Alus was higher within DM uniquely overexpressed Alus (median 10.6%, IQR 6.4%-15.9%). Alterations in overall Alu expression and/or Pol III transcribed Alus can be caused by altered activity of DNA Methyltransferase enzymes, but we observed no clear alteration in their expression that was specific to DM (**fig. S1**)

Alus are divided into three subfamilies (J, S, and Y), based on their insertional age into the human genome, with the Y subfamily being the youngest.[12] Older repeat families usually accumulate mutations improving read alignment since their transcripts become more unique from each other. To explore a potential bias of undercounting the younger elements, we quantified the average per sample expression of Alus by their subfamilies and found no decrease in expression of the Y subfamily compared to the two other subfamilies (**fig. S2**), indicating no underestimation in the expression of the youngest Alu subfamily due to multi-mapping.

### A subset of uniquely expressed Alus in DM correlated with interferon stimulated genes and with markers of disease activity

We correlated expression of the top 10 elements with lowest q-values among uniquely expressed Alus with the average expression of type I interferon-stimulated genes (ISGs) in myositis subtypes. As shown in **Fig. 3a**, Alu transcripts in DM showed the highest correlation with ISGs (Spearman r=0.9, q= 8.8e-16), compared to AS (r=0.56, p=0.041), IMNM with a negative correlation, and IBM that showed no statistically significant correlation. Next, we examined the top 5 Alus most correlated with ISGs (**Fig. 3b**), four of which were also among the top 10 uniquely overexpressed in DM.

**Fig. 3:**
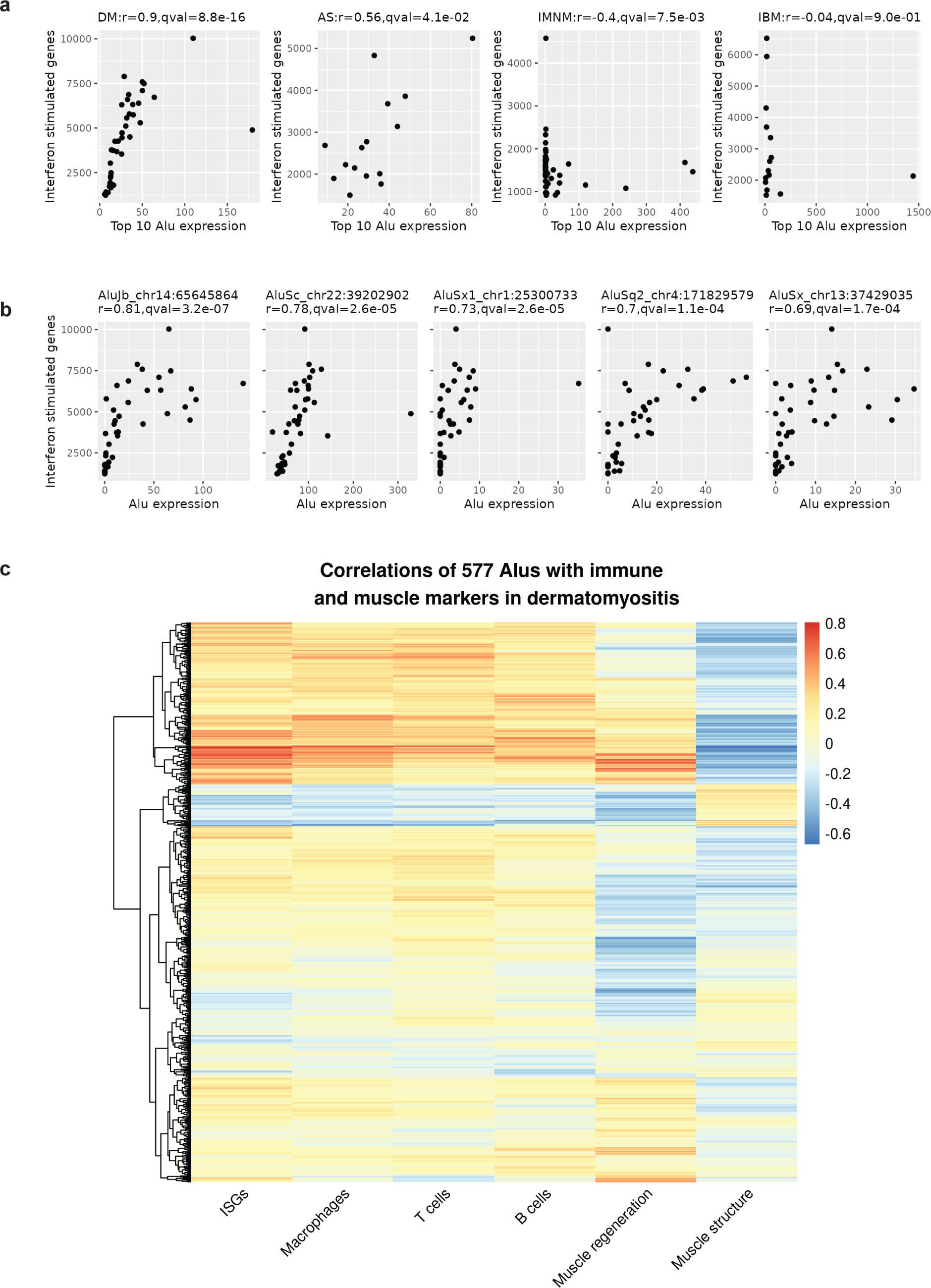
Correlation of Alus with interferon stimulated genes and disease markers in myositis. **a**, Spearman correlations between the top 10 Alus with lowest q values and interferon-stimulated genes (ISGs) in the four myositis subtypes. **b**, Spearman correlations between the 5 individual Alus most correlated with interferon-stimulated genes (ISGs) in dermatomyositis. **c**, Heat map of Spearman correlations between the 577 overexpressed Alu elements with ISGs and gene markers of macrophages, T and B cells, muscle regeneration and muscle structure.

While a majority of the uniquely overexpressed Alus in DM did not correlate with ISGs, a subset of 167 Alus clustered together showing high correlations with ISGs as well as gene markers for macrophages, T cells, B cells, and muscle regeneration and negatively with muscle structural gene expression (**Fig. 3c**), indicating these as more likely to be relevant in disease biology. This list included 102 (61.1%) elements from the Alu S family, 43 (25.7%) from Alu J, and 19 (11.4%) from the Alu Y family. The disease-associated Alus included 128 (76.6%) intergenic/antisense, 33 (19.8%) intronic, and 6 (3.6%) exonic elements, that again clustered on chromosome 19 (n=22, 13.2%) and in a pericentromeric region of chromosome 11. Of these Alus on chromosome 19, four were intronic inside *DNAAF3* (a dynein assembly gene), *ZNF497*, *AC092296.2*, and *AC130469.1*.

### Alu A-to-I RNA editing

A-to-I RNA editing by ADAR creates breaks in dsRNA preventing it from triggering cellular dsRNA sensors if sufficiently edited. Alu transcripts are the predominant targets of ADAR. Therefore, we quantified A-to-I RNA editing and found 16,277 distinct edited adenosines in 6,417 Alu elements. This contrasts with RNA editing in other genomic repeats (n=544 editing sites in 133 repeats). We illustrated an example of an edited Alu on chromosome 16 (**Fig. 4a**) embedded inside a LINE2 element, with a nearby reverse complementary Alu. This lies in a region of the genome that was recently deciphered by the T2T genome.

**Fig. 4:**
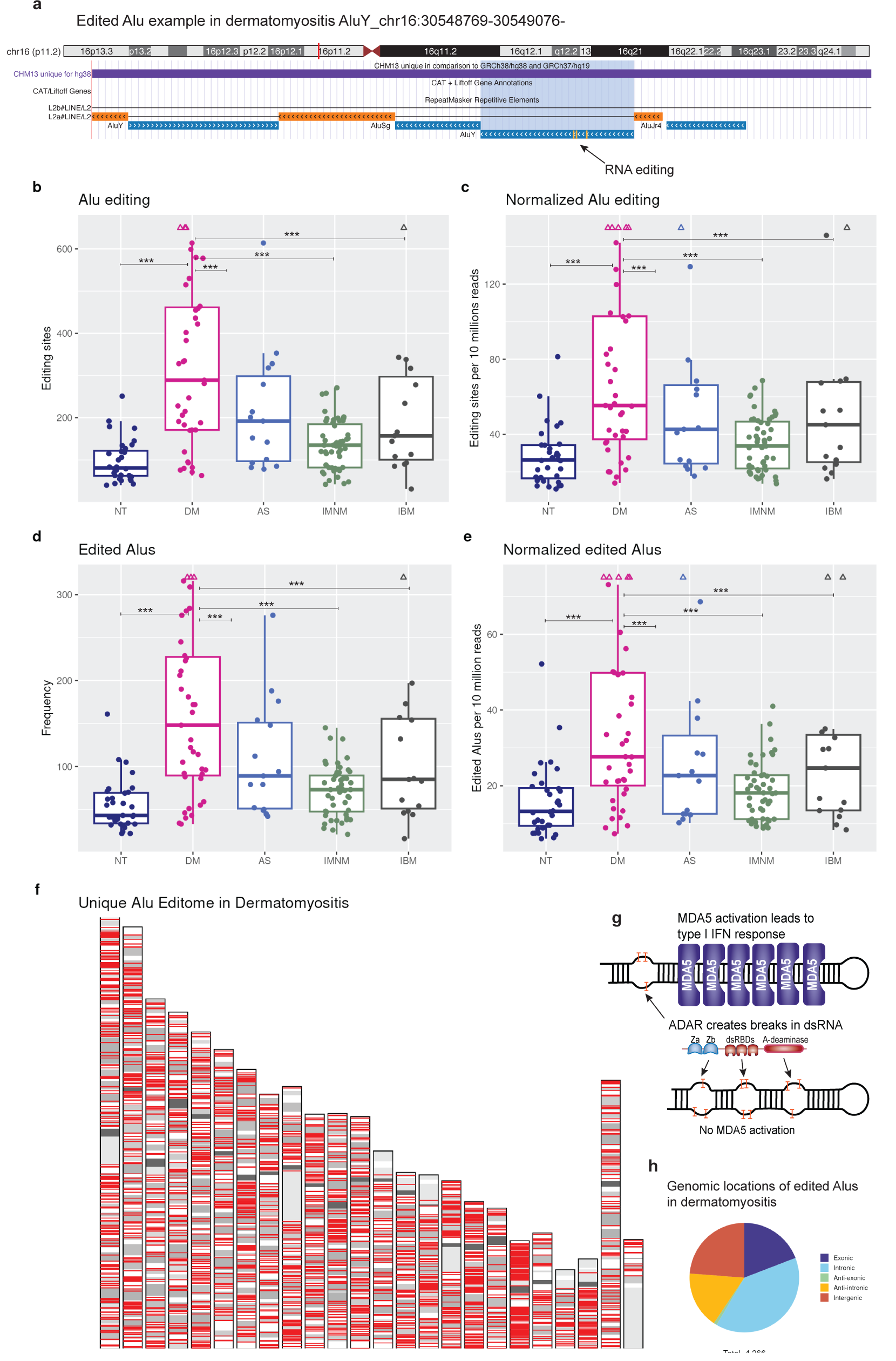
Alu A-to-I RNA editing in myositis. **a**, Illustrative example of an individual A-to-I edited Alu in DM in a region of the genome that is unique to the T2T genome. **b-e**, Raw counts and normalized counts by sequencing depth of Alu A-to-I editing sites and edited Alus in normal muscle and myositis subtypes, Poisson regression, ***= q<0.001. **f**, Chromosomal location (red lines, not to scale) of the edited Alus in dermatomyositis not seen in normal muscle or other myositis types. **g**, Schematic illustration of dsRNA editing by ADAR with the domain structure of ADAR, long dsRNA that is either unedited or insufficiently edited will trigger MDA5 by oligomerization of adjacent MDA5 molecules. **h**, Genomic location of edited Alus in dermatomyositis with respect to genes. For all boxplots, center line, median; box limits, upper and lower quartiles; whiskers, 1.5x interquartile range; points beyond whiskers, outliers (triangles when higher than maximum whisker).

Alu RNA editing was increased in DM compared to normal tissue and other myositis types. The median number of Alu editing sites per sample (**Fig. 4b**) was highest in DM (289, interquartile range [IQR] 171-462), compared to healthy muscle (81, IQR 62.2-122, p<0.001), AS (192, IQR 97-298, p<0.001), IMNM (135, IQR 82-184, p<0.001), and IBM (157, IQR 100-298, p <0.001). To account for sequencing depth, we also quantified Alu editing sites per 10 million sequencing reads and found similar results (**Fig. 4c**). In addition to counting A-to-I editing sites, we counted distinct edited Alus per sample and again saw similar patterns where DM had the highest numbers of edited Alus (**Fig. 4 d-e**).

The number of Alu transcripts that were uniquely edited in DM was higher (n=2,996) than in normal muscle (n=226), or in the other myositis subtypes AS (n=356), IMNM (n=539), and IBM (n=870). To better illustrate that other myositis subtypes had fewer uniquely edited Alu transcripts, we repeated this calculation excluding DM and found similarly lower numbers of unique edited Alus in NT (n=298), AS (n=457), IMNM (n=723), and IBM (n=1098). Uniquely edited Alus in DM showed widespread distribution in all chromosomes (**Fig. 4f**) with clustering in chromosome 19 (n=193), which had the 3^rd^ highest number of edited Alus after chromosomes 1 (n=258) and 2 (n=214), despite being about four times smaller in size.

Because ADAR only edits dsRNA, observed A-to-I RNA editing indicates the presence of dsRNA in vivo[13, 14] (illustrated in **Fig. 4g**), allowing construction of editomes that reflect dsRNA. The clustering of Alu expression and editing in chromosome 19 likely reflects a known high concentration of dsRNA in chromosome 19.[14] The majority of edited Alus in DM were either intronic (39.3%) or exonic (19.2%), see **Fig. 4h**, which is consistent with prior studies of MDA5-protected dsRNA.[11] We show the complete editomes of myositis types and healthy muscle in **Fig. 5 a-e.**

**Fig. 5:**
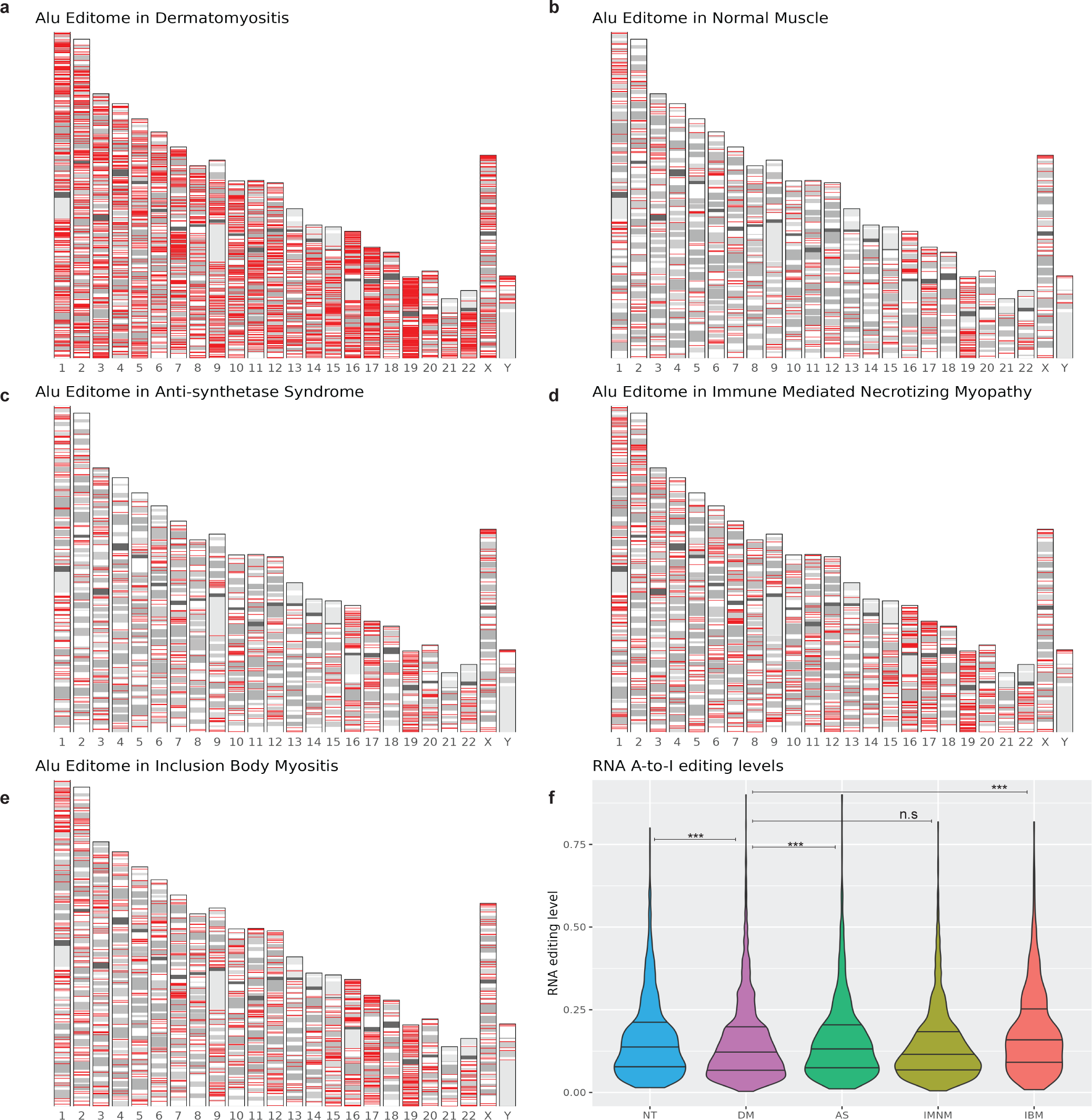
Alu A-to-I editomes and editing levels in myositis. **a-e**, Chromosomal locations (red lines, not to scale) of edited Alus in myositis types and normal muscle. **f**, Alu editing levels (number of reads with editing divided by total read coverage at site), included sites with coverage of ≥10 reads, logistic regression, ***= q<0.001, n.s.= not statistically significant.

Increases in numbers of editing sites can be due to increased dsRNA or ADAR activity/capacity. The expression of *ADAR*, but not the related *ADARB1*, was increased in DM muscle (**fig. S3)**, which is not unexpected since it is an interferon stimulated gene. However, it has been reported that *ADAR* mRNA levels are not good predictors of editing levels.[15] If the editing activity/capacity was increased, we’d expect to see an increase in the percent of edited reads out of total reads covering a site (editing level). Therefore, we quantified editing levels by myositis group and found that DM had a higher percent of low-level editing (16.8%), compared NT (14.4%) and other myositis types (range 11.5%-15.5%). Similarly, the distribution of editing levels at RNA editing sites were either lower or unchanged in DM compared to other myositis and normal tissue (**Fig. 5f**). We conclude that the observed increase in editing sites and edited Alus in DM cannot be explained by an increase in editing activity/capacity alone.

### A-to-I RNA editing inside genes

In addition to Alu editing, we also detected A-to-I editing of transcripts other than Alus, including inside genes (n= 6,491 A-to-I edits). However, the majority were antisense to Alu elements (n=4828, 74%) and showed similar expression pattern to Alu editing (**Fig. 6a**). These likely represent editing of an dsRNA formed by an Alu element inside the gene with a reverse complementary sequence of an Alu from the antisense strand.

**Fig. 6:**
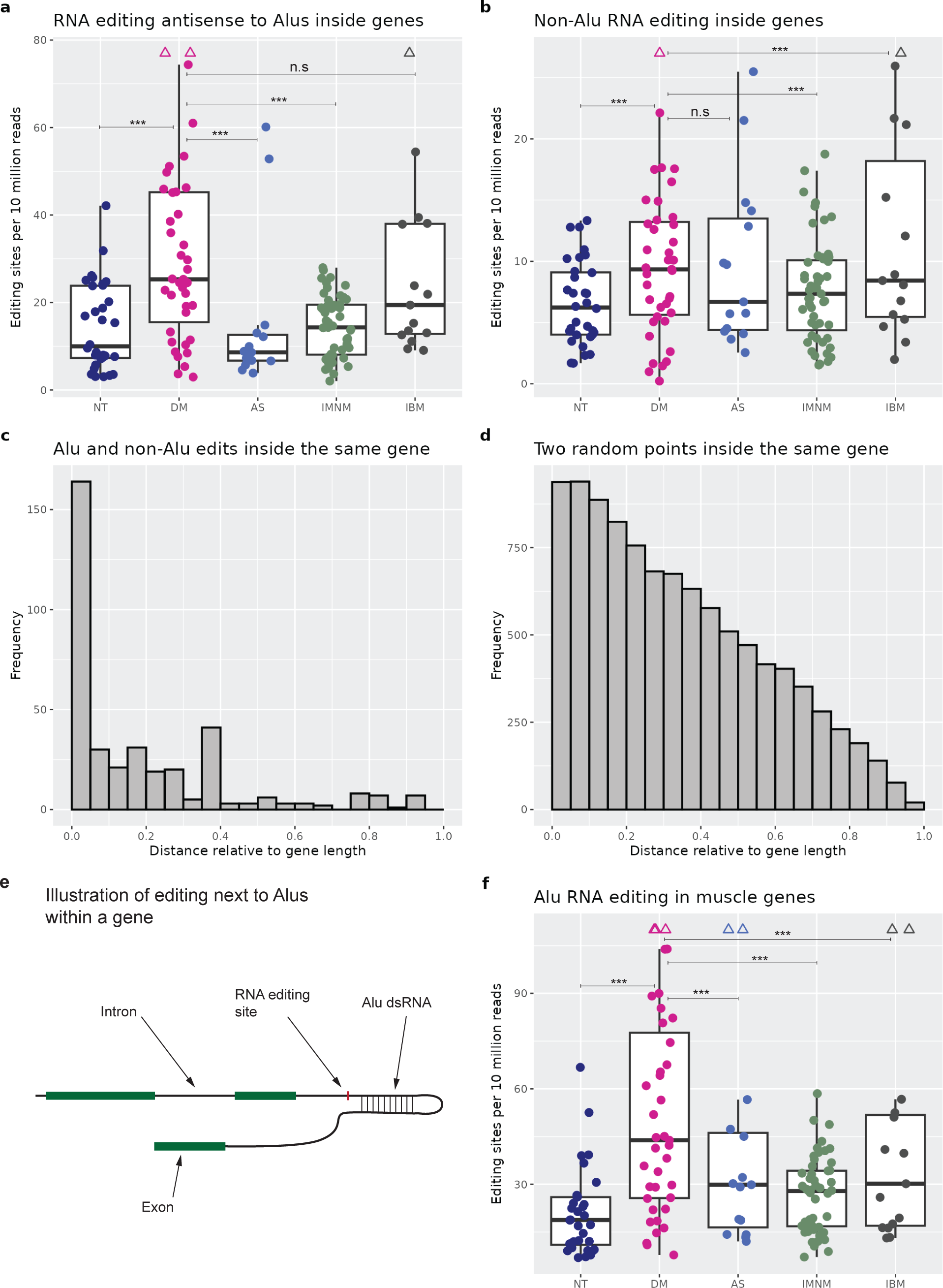
A-to-I RNA editing inside genes. **a**, Editing sites inside genes antisense to Alus in normal muscle and the myositis subtypes, Poisson regression, ***= q<0.001, n.s.= not statistically significant. **b**, Non-Alu editing sites inside genes in normal muscle and the myositis subtypes, Poisson regression, ***= q<0.001, n.s.= not statistically significant. **c**, Distance between non-Alu editing sites and Alu sequences inside genes (n= 374). **d**, Distance between two randomly selected nucleotides in hypothetical genes (n=10,000 permutations). **e**, Illustration of non-Alu editing inside a gene nearby to dsRNA formed by Alus. **f**, Counts of Alu editing sites inside muscle specific genes, Poisson regression, ***= q<0.001, n.s.= not statistically significant. For all boxplots, center line, median; box limits, upper and lower quartiles; whiskers, 1.5x interquartile range; points beyond whiskers, outliers (triangles when higher than maximum whisker).

Excluding editing in Alus and antisense to Alus, there were 1,663 editing sites inside genes, see **Fig. 6b**, with 374 (22.5%) of these edits being in transcripts that also contained edited Alus. The median distance between non-Alu edits and same-gene Alus was 6,278 bases (IQR 1038-30,704). Relative to gene length, the edited Alus and non-Alu edits were within a median distance of 7.0% apart (IQR 3.3%-29.6%), see **Fig. 6c**, compared to 2 randomly picked points in hypothetical genes of varying lengths (**Fig. 6d**). The large difference between these two graphs indicates that edited Alus and same-gene non-Alu edits are much closer to each other than what is expected by chance. The dsRNA formed by Alus in these genes likely facilitated editing of flanking adenosines in nearby non-Alu sequences, a phenomenon that has been described before,[16] illustrated in **Fig. 6e**.

Muscle biopsies contain multiple types of cells. In order to investigate whether RNA editing was occurring inside muscle fibers, we selected a group of genes from a muscle contraction pathway as well as the most highly expressed genes in skeletal muscle from the GTEx project, likely reflecting more muscle specific genes. RNA editing inside these genes showed a similar pattern with increased editing in DM (**Fig. 6f**) providing a clue that the observed patterns of editing included muscle fibers at least partially.

## DISCUSSION

In this study, we report that DM is characterized by overexpression of hundreds of Alus in a unique way not seen in other myositis types. A subset of these Alus correlated with interferon stimulated genes, lymphocytes, macrophages, and other markers of disease activity. In addition, Alu transcripts with evidence of A-to-I editing (reflecting dsRNA) were also increased in DM more than in other myositis types. Based on these results, we introduce a new hypothesis in DM pathogenesis that increased Alus create excessive dsRNA potentially triggering MDA5 and interferon production.

The cause of the Alu overexpression in DM remains unknown. Interferons have been reported to stimulate Alu transcription.[17] However, if interferon caused the large and unique pattern of Alu element expression we observe in DM, we would expect that all these Alus would correlate closely with the ISGs. This is not the case, the majority of the uniquely overexpressed Alus in DM did not correlate with ISGs, suggesting a different mechanism of induction, such an epigenetic dysregulation over large areas of the genome. Further research is needed to clarify the cause(s) of Alu overexpression in DM.

Our results indicate that there was an increase in edited Alus in DM more than what is seen in normal muscle or other myositis types. Multiple studies have shown that skeletal muscle exhibits the lowest levels of A-to-I editing compared to all other studied tissues.[15, 18] Hence, muscle tissue might be less tolerant of increased endogenous dsRNA. Because ADAR only edits dsRNA,[13, 14] the increased Alu editing means higher levels of Alu dsRNA in DM. Based on this we hypothesize that Alu overexpression creates excessive amounts of endogenous dsRNA that may overwhelm the editing capacity of ADAR, leading to excessive levels of unedited or insufficiently edited Alu dsRNA that trigger RNA sensors and interferon production. An increase in Alu dsRNA is by itself sufficient to trigger MDA5. This has been shown in prior research of demethylating agents in cancer causing an increase in Alu expression overwhelming the editing capacity of ADAR and leading to interferon production.[11]

Our use of the T2T reference genome for alignment is a strength for our study because it allows more accurate identification of repeat elements. For example, we observed editing of an intergenic Alu on chromosome 16 (p11.2) in a region of the genome that is visible in T2T, but not in reference genome Hg38. Nevertheless, quantification of transposable elements from RNA sequencing data is challenging. Due to their abundance and similarity, there will be reads that map equally well to multiple elements (multi-mappers). If these are excluded, there is a risk of biasing results towards older elements that had more evolutionary time to accumulate more mutations making them more unique and resulting in less multimapping. On the other hand, there is no ideal method to deal with multi-mappers, some bioinformatics tools will distribute them randomly or use other methods.[19] However, these cannot completely circumvent the uncertainty of multi-mappers and introduce other problems, e.g. an element that is not expressed at all will appear to be expressed if reads arising from different elements shared multi-mappers. We decided it was more important to avoid counting non-existent elements and focused on elements that we can be confident were in fact expressed. Therefore, we used only uniquely mapped reads. In addition, because SINEs are much shorter than other transposable elements such as LINEs and HERVs, multimapping is likely less of an issue. To support the absence of this type of bias in our results, we quantified Alu expression by evolutionary family (Y, S, J), and found that the youngest family (Y) did not have lower expression compared to the S and J (oldest) families.

In summary, we show that increased Alus and RNA editing occur in DM, in a pattern not seen in other myositis types, despite these diseases having similar and overlapping clinical features and histological features. Future research will be needed to confirm the exact identity of dsRNA bound to RNA sensors in DM and to identify the causes of increased Alu expression in DM. Because autoantigens in dermatomyositis exhibit RNA binding properties including MDA5,[9] NXP2,[20] TIF1g,[21] and Mi2,[22] our results open the doors to additional research avenues to explore if myositis specific autoantibodies form due to autoantigens binding to or being in complex with[23] immunogenic dsRNA.

## METHODS

### RNA sequencing data

We used publicly available bulk RNA sequencing data of distinct muscle biopsies from patients with idiopathic inflammatory myopathies, n=165 (GEO accession GSE220915). These data included stranded single-end 50 bp reads. Since we have used these data before,[24] we depended on our prior QC measures, and excluded 13 samples due to relatively lower quality (poor gene body coverage and/or high proportions of unmapped reads). The remaining 152 samples were analyzed by myositis subgroups. We used STAR[25] (v. 2.7.11b) to align reads to the T2T reference genome (CHM13v2.0), which is the first truly complete human reference genome allowing more accurate identification of transposable elements.[3]

### Identification of short interspersed elements

Short interspersed elements (SINEs) were identified using BEDTools[26] (v. 2.31.1), by intersecting T2T SINE coordinates with aligned reads, with a requirement to be on the same strand. Spliced reads were handled with the BEDTools “-split” option to avoid intersecting gaps between spliced reads. A matrix of SINE frequencies per sample was constructed using only uniquely mapped reads to avoid uncertainty from multimapping reads.

### Differential expression of SINEs

SINEs with ≥10 reads in ≥4 samples were included and tested for differentially expression of each myositis group vs normal muscle tissue using DESeq2 (v. 1.38.3).[27] Statistical significance was set at an alpha=0.05 for adjusted p values (q values). To find SINEs uniquely expressed in one group and not in other myositis types or normal muscle, we start with the differentially expressed SINEs for each group, then subtract elements with same-direction expression (vs control) with q values <0.2 in any other group. This higher q value cutoff is more conservative resulting in a smaller set of uniquely expressed elements. Finally, we focus on elements with ≥2 fold change vs controls.

### Genomic location of SINEs

T2T CHM13v2.0 cat/liftoff gene and exon coordinates were downloaded from the UCSC Table Browser[28] on August 6, 2024, and were intersected with the identified SINEs to classify them into exonic or intronic. SINEs located on opposite strands to exons or introns were classified as anti-exonic or anti-intronic, respectively. Finally, SINEs outside these locations were classified as intergenic. Since an element may fit more than one category, e.g. an element can be inside an intron and on opposite strand to a different gene, the groups were made mutually exclusive based on the following order: exonic, intronic, anti-exonic, anti-intronic, and intergenic.

### Gene expression

Gene transcript quantification was performed as previously described[24] followed by differential expression at the gene level for each myositis group vs normal tissue using DESeq2. Genes with ≥10 reads in ≥4 samples were included. Average expression of gene markers of interest was quantified per sample. This included type 1 interferon stimulated genes (ISGs) based on a list from the GSEA hallmark pathway,[29] Markers for macrophages (*CD14*, *CD68*), T cells (*CD3D*, *CD3E*, *CD4*, *CD8A*), B cells (*CD79A*, *CD79B*), muscle regeneration (*NCAM1*, *MYOG*, *MYOD1*, *PAX7*), and adult muscle structural proteins (*ACTA1*, *MYH1*, *MYH2*).

### RNA pol III transcription

Alus can be transcribed passively by Pol II when they’re inside genes, or they can be transcribed by Pol III through their own promoters. We detected Pol III transcription by quantifying RNA sequencing coverage at the Alu element itself and in the areas upstream and downstream. Alus with ≥10 read coverage were included. The Alu body coverage area was defined to start 5 bases upstream and 20 bases downstream of the Alu element. Using established paramters,[6] we applied a cutoff for the upstream segment to have less than 1/7 the coverage of the Alu body, while the downstream segment had to have no more than half the Alu body coverage. This is because Alu transcription may continue till encountering a stop codon potentially extending beyond the Alu element.

### RNA editing

A-to-I RNA editing was quantified using SPRINT[30] (v. 0.1.8) which implements SNP-free methodology based on clustering patterns of single nucleotide variants duplets. A-to-I editing is detected as A-to-G, which appears as T-to-C editing when occurring on the antisense strand.[30, 31] Therefore, we included sense A-to-G and antisense T-to-C editing to represent the full spectrum of A-to-I RNA editing. RNA editing can be extensive resulting in unmapped reads, referred to as “hyper editing”. We quantified both regular and hyper editing and included only editing sites with ≥2 read support as described by SPRINT methodology. The first six bases of each read were ignored since these commonly include high technical variation due to using random hexamers primers during RNA-sequencing library preparation. Alu RNA editing was identified by intersecting genomic coordinates of editing sites with that of T2T Alus.

### Statistical testing

Data management and statistical analyses were performed using R (v. 4.2.1).[32] Proportions were tested using the binomial test. Correlation coefficients were calculated using the Spearman method. Count data were modelled with Poisson regression, and binary outcomes were modelled with regression models. All p values were two-sided and were adjusted for multiple comparisons using the Benjamini–Hochberg method.[33] Statistical significance was set at alpha=0.05.

## Data Availability

All data are available online at https://www.ncbi.nlm.nih.gov/geo/query/acc.cgi?acc=GSE220915

## Acknowledgements

We thank Dr. Iago Pinal-Fernandez for his help in acquiring the data.

## Author contributions

RN conceptualized the research project. AM acquired the data. RN carried out the analysis. RN and TM interpreted the results and planned additional analyses as appropriate, with help from AM. RN wrote the first draft of the manuscript. All authors contributed to reviewing and editing the manuscript. All authors provided pertinent feedback that improved the quality of the manuscript.

## Supplemental figures

**Fig. S1:**
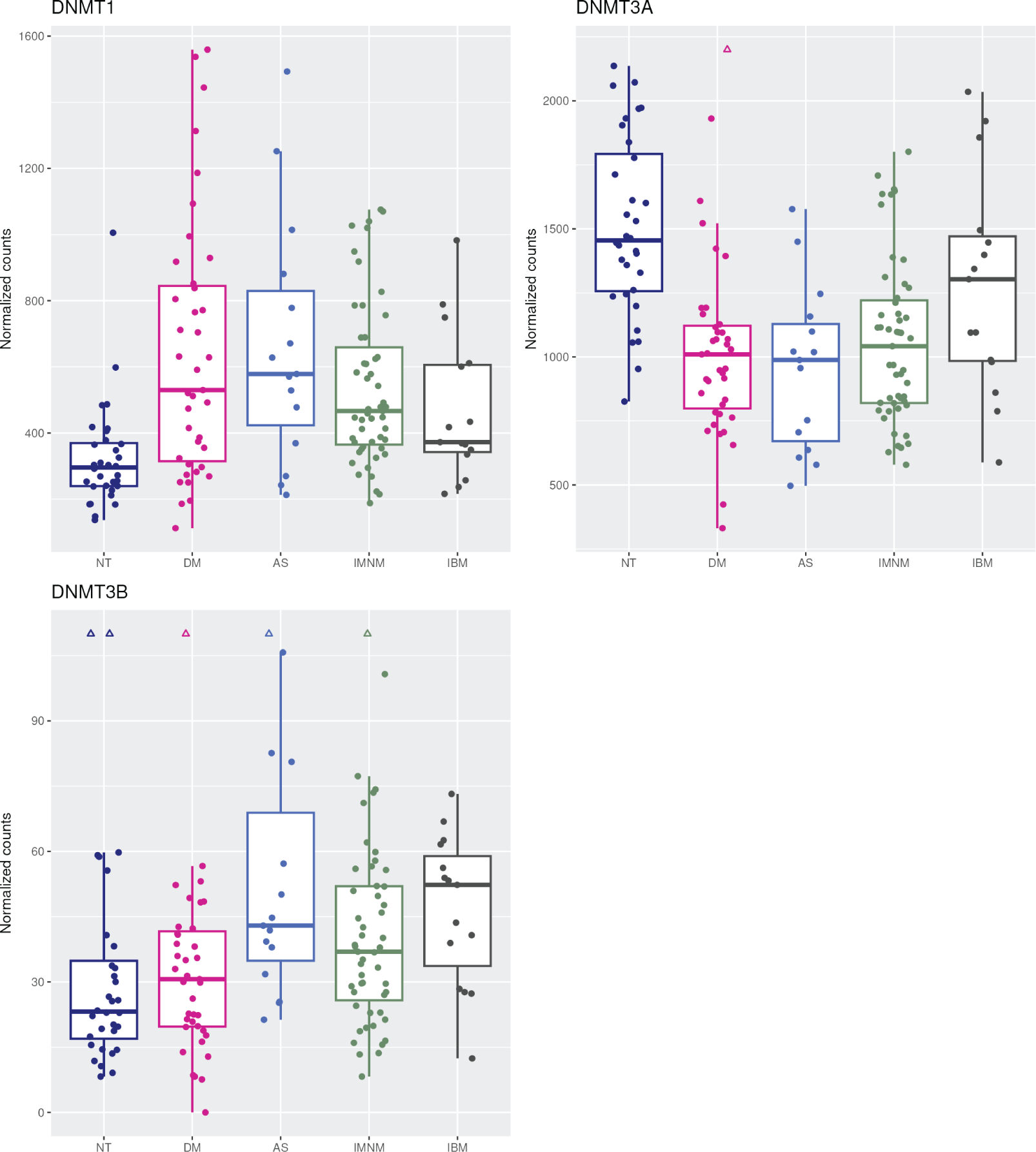
Expression of DNMT genes in myositis.

**Fig. S2:**
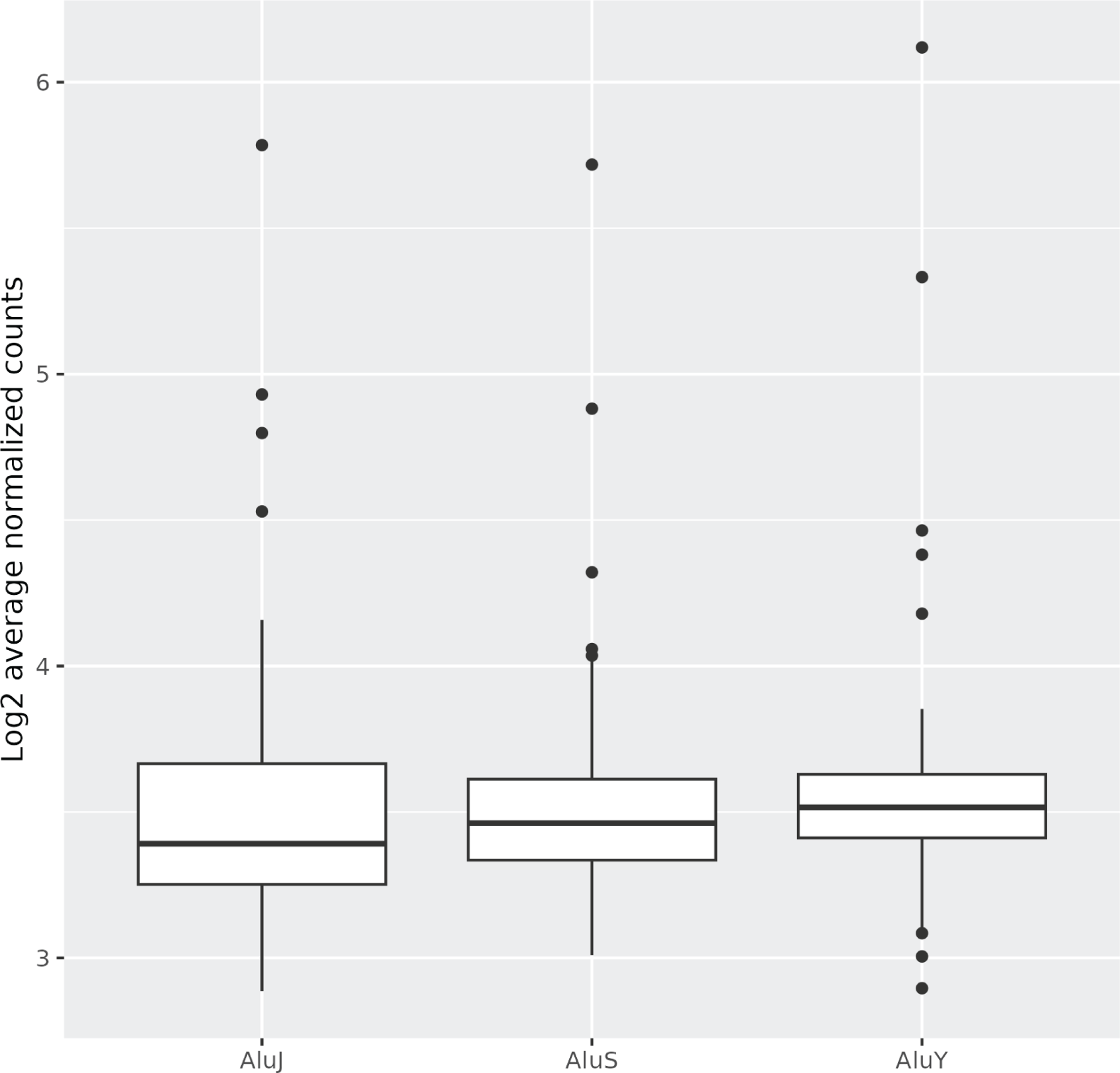
Expression of Alu subfamilies in myositis.

**Fig. S3:**
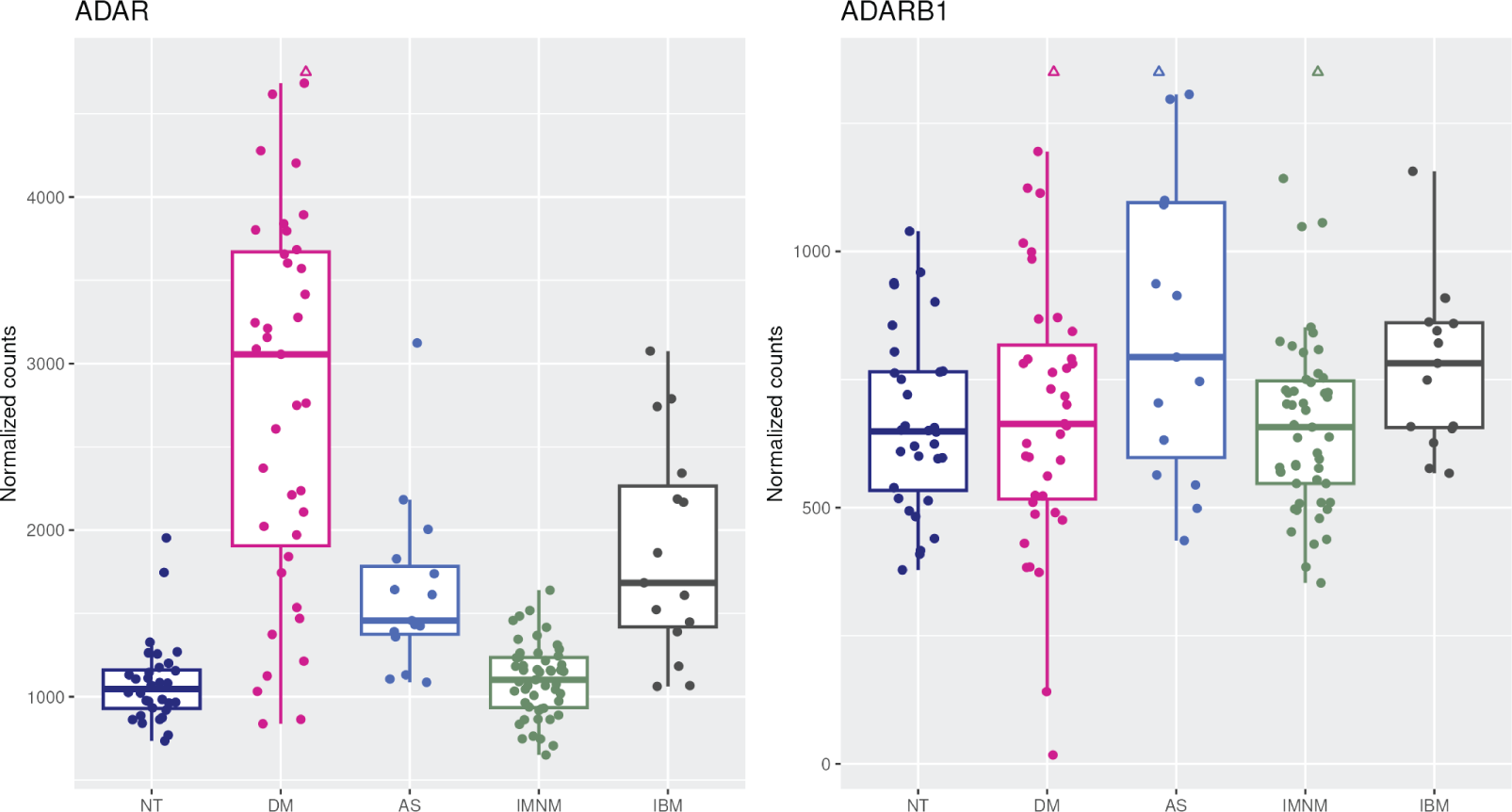
Expression of ADAR genes in myositis.

## References

1. Pinal-Fernandez, I., et al., Identification of distinctive interferon gene signatures in di2erent types of myositis. Neurology, 2019. 93(12): p. e1193–e1204.

2. Lundberg, I.E., M. de Visser, and V.P. Werth, Classification of myositis. Nat Rev Rheumatol, 2018. 14(5): p. 269–278.

3. Hoyt, S.J., et al., From telomere to telomere: The transcriptional and epigenetic state of human repeat elements. Science, 2022. 376(6588): p. eabk3112.

4. Lander, E.S., et al., Initial sequencing and analysis of the human genome. Nature, 2001. 409(6822): p. 860-921.

5. Zhang, X.O., H. Pratt, and Z. Weng, Investigating the Potential Roles of SINEs in the Human Genome. Annu Rev Genomics Hum Genet, 2021. 22: p. 199–218.

6. Conti, A., et al., Identification of RNA polymerase III-transcribed Alu loci by computational screening of RNA-Seq data. Nucleic Acids Res, 2015. 43(2): p. 817–35.

7. Carnevali, D., et al., Whole-genome expression analysis of mammalian-wide interspersed repeat elements in human cell lines. DNA Res, 2017. 24(1): p. 59–69.

8. Kim, S., et al., Evidence of Aberrant Immune Response by Endogenous Double-Stranded RNAs: Attack from Within. Bioessays, 2019. 41(7): p. e1900023.

9. Ahmad, S., et al., Breaching Self-Tolerance to Alu Duplex RNA Underlies MDA5-Mediated Inflammation. Cell, 2018. 172(4): p. 797–810 e13.

10. Bahn, J.H., et al., Genomic analysis of ADAR1 binding and its involvement in multiple RNA processing pathways. Nat Commun, 2015. 6: p. 6355.

11. Mehdipour, P., et al., Epigenetic therapy induces transcription of inverted SINEs and ADAR1 dependency. Nature, 2020. 588(7836): p. 169-173.

12. Deininger, P., Alu elements: know the SINEs. Genome Biol, 2011. 12(12): p. 236.

13. Sijen, T. and R.H. Plasterk, Transposon silencing in the Caenorhabditis elegans germ line by natural RNAi. Nature, 2003. 426(6964): p. 310-4.

14. Reich, D.P. and B.L. Bass, Mapping the dsRNA World. Cold Spring Harb Perspect Biol, 2019. 11(3).

15. Roth, S.H., E.Y. Levanon, and E. Eisenberg, Genome-wide quantification of ADAR adenosine-to-inosine RNA editing activity. Nat Methods, 2019. 16(11): p. 1131–1138.

16. Ramaswami, G., et al., Accurate identification of human Alu and non-Alu RNA editing sites. Nat Methods, 2012. 9(6): p. 579–81.

17. Hung, T., et al., The Ro60 autoantigen binds endogenous retroelements and regulates inflammatory gene expression. Science, 2015. 350(6259): p. 455-9.

18. Gabay, O., et al., Landscape of adenosine-to-inosine RNA recoding across human tissues. Nat Commun, 2022. 13(1): p. 1184.

19. Lanciano, S. and G. Cristofari, Measuring and interpreting transposable element expression. Nat Rev Genet, 2020. 21(12): p. 721–736.

20. Kimura, Y., et al., The newly identified human nuclear protein NXP-2 possesses three distinct domains, the nuclear matrix-binding, RNA-binding, and coiled-coil domains. J Biol Chem, 2002. 277(23): p. 20611–7.

21. Weng, L., et al., The E3 ubiquitin ligase tripartite motif 33 is essential for cytosolic RNA-induced NLRP3 inflammasome activation. J Immunol, 2014. 193(7): p. 3676–82.

22. Ullah, I., et al., RNA inhibits dMi-2/CHD4 chromatin binding and nucleosome remodeling. Cell Rep, 2022. 39(9): p. 110895.

23. Sanderson, N.S., et al., Cocapture of cognate and bystander antigens can activate autoreactive B cells. Proc Natl Acad Sci U S A, 2017. 114(4): p. 734–739.

24. Najjar, R., et al., Distinct Transcript-Level Expression Profiles and Unique Alternative Splicing in Inflammatory Myopathies. ACR Open Rheumatol, 2024.

25. Dobin, A., et al., STAR: ultrafast universal RNA-seq aligner. Bioinformatics, 2013. 29(1): p. 15–21.

26. Quinlan, A.R. and I.M. Hall, BEDTools: a flexible suite of utilities for comparing genomic features. Bioinformatics, 2010. 26(6): p. 841–2.

27. Love, M.I., W. Huber, and S. Anders, Moderated estimation of fold change and dispersion for RNA-seq data with DESeq2. Genome Biol, 2014. 15(12): p. 550.

28. Nassar, L.R., et al., The UCSC Genome Browser database: 2023 update. Nucleic Acids Res, 2023. 51(D1): p. D1188–D1195.

29. Subramanian, A., et al., Gene set enrichment analysis: a knowledge-based approach for interpreting genome-wide expression profiles. Proc Natl Acad Sci U S A, 2005. 102(43): p. 15545–50.

30. Zhang, F., et al., SPRINT: an SNP-free toolkit for identifying RNA editing sites. Bioinformatics, 2017. 33(22): p. 3538–3548.

31. Pecori, R., et al., ADAR RNA editing on antisense RNAs results in apparent U-to-C base changes on overlapping sense transcripts. Front Cell Dev Biol, 2022. 10: p. 1080626.

32. Team, R.C., R: A language and environment for statistical computing. R Foundation for Statistical Computing, Vienna, Austria, 2022.

33. Benjamini, Y. and Y. Hochberg, Controlling the False Discovery Rate: A Practical and Powerful Approach to Multiple Testing. Journal of the Royal Statistical Society: Series B (Methodological), 1995. 57(1): p. 289–300.

